# A Multidisciplinary Approach: Teaching Medical Spanish to Medical Students Using Role-play

**DOI:** 10.1101/2020.10.19.20214809

**Authors:** Khashayar Mozaffari, Rebecca Kolodner, Eric Chalif, Victor Valdivia Ruiz, Benjamin Blatt

## Abstract

**Introduction:** The Hispanic community is the most rapidly growing minority group in the United States, making up 18.1% of the population, with 40% reporting limited English proficiency. To address this need, many health sciences institutions have implemented medical Spanish courses to increase Spanish proficiency among future healthcare providers. Although interactive courses have shown efficacy in teaching field-related terminology, barriers to medical Spanish curriculum implementation persist. Our study investigated the benefit of role-play, an underutilized teaching modality, in a medical Spanish course.

**Methods:** 19 second year medical students were recruited to participate. Based on their placement test performance, students were assigned to a beginner or intermediate group and met weekly for one-hour sessions over five consecutive weeks. Students assumed the roles of Spanish-speaking patient, English-speaking provider, and interpreter to practice various medical scenarios. Students completed pre- and post-course examinations to assess Spanish proficiency improvement. Statistical significance was determined using a *p*-value < 0.05.

**Results:** Seven students, all members of the intermediate group, completed the course. Attendance among this group was 77.4%. When comparing examination scores, there was statistically significant improvement in oral translation of phrases from Spanish to English (*p*-value= 0.03).

**Discussion:** Statistically significant improvement in oral translation of phrases from Spanish to English was accomplished through a minimal time requirement of one hour per week utilizing role-play. Given the fact that limited time poses a barrier to implementing medical Spanish curricula, our findings highlight the potential benefit of this teaching methodology and call for further evaluation.

## Introduction

There are over 59.9 million Hispanics in the United States [1]. This number makes up about 18.3% of the nation’s total population and is expected to increase to 24.5% by 2050 [1]. Hispanics are the most rapidly growing minority group in the country, and nearly 40% of Hispanic patients are categorized as having limited proficiency in English [2]. Such numbers draw attention to the nationwide shortage of certified medical interpreters and the need for Spanish-speaking providers in the health care system [3]. Language concordance is a pivotal component of physician-patient relationship and it leads to higher patient satisfaction and better long-term outcomes [4–10]. With the aim of training future healthcare providers with adequate proficiency in Spanish, many health sciences institutions in the United States have implemented medical Spanish courses. Although interactive classes have shown to be effective in teaching field related terminology to students, there are still significant barriers to implementing a medical Spanish curriculum [2,11–13]. According to a national survey of medical schools in the United States, lack of time is the most frequently reported barrier to implementing a medical Spanish course [2]. In addition, there is a sparsity of well-established assessment methods to evaluate the effectiveness of such courses [12,14–16]. Also, healthcare providers tend to overestimate their proficiency in the language, and thus it is important to develop an objective assessment to accurately evaluate their competency and also the effectiveness of the course [17–19]. The potential collaboration with language instructors can help provide a more comprehensive course which has shown to be beneficial [14,18,20]. This can also add an emphasis on cultural competency to the objectives of the class as language instructors help integrate such aspects to the course. Being culturally competent in addition to having conversational Spanish skills leads to more satisfactory patient outcomes [21–23]. We utilized a collaborative approach with the Spanish department faculty to effectively design an interactive course that utilized role-play as the main teaching method with focus on medical knowledge and cultural aptitude. Role-play has been suggested as minimum best practice as it offers an interactive component essential to patient care [24]. Despite this, there is great variability in role-play simulation across programs. This modality, originally borrowed from the language acquisition setting, has been widely used to teach effective communication skills to medical students and residents alike [25]. Despite evidence that role-play can be used across varied learners, content and under different time-constraints, only 2/3rds of programs surveyed nationally reported using student-to-student role-play in their medical Spanish curricula [26]. The goal of this study was to evaluate the potential of role-play interaction as a time efficient method of improving comprehension of Spanish language field related terminology, communication skills, and cultural competency of beginner to intermediate proficiency students. This article aims to address the gap in the current literature on time-efficient teaching modalities such as role-play in medical student education, and promote its further exploration and utilization in medical Spanish curricula.

## Methods

In a study reviewed and approved by the Institutional Review Board (NCR191414), incoming second-year medical students were recruited to participate via class-wide emails and verbal announcements during classes at George Washington University School of Medicine and Health Sciences (GWSMHS). GWSMHS matriculated an entering class of 185 medical students. Prior to course participation, 19 second year medical students were informed about the goals of the study and reminded that their participation was voluntary. All participants provided their informed consent. The course material was designed by a Spanish-fluent medical student with experience working as a certified medical interpreter, and a Spanish professor in the language department of the university. On the first day of the course, students were given a placement exam containing 46 total questions composed of oral and multiple-choice components. The exam was designed by the course instructor and approved by the Spanish department faculty. The multiple-choice portion aimed to test the students’ knowledge of the terminology, while the oral portion tested the same concepts in addition to evaluating the students’ ability to verbally translate phrases from English to Spanish and vice versa. Such questions evaluated the students’ knowledge of general vocabulary and ability to converse (Appendix). The exam was proctored and later graded by the course instructor. Based on their performances, students were assigned to either a beginner or intermediate group. The classes held one-hour weekly sessions for five consecutive weeks. Students received a copy of the upcoming session’s vocabulary lesson and practice scenario a few days before class via email and were encouraged to review them prior to coming to class. The vocabulary lesson contained medical-related terminology including names of body organs such as the liver, lungs, heart, kidneys, etc., and also health-care related words such as physician, nurse, social worker, etc. Each class was structured in the following way: The first twenty minutes of the session were dedicated to learning relevant vocabulary pertinent to that session. The objective here was to introduce the words that students would encounter in the session’s role-play scenario. Subsequently, the instructor introduced a role-play scenario to students with settings including inpatient hospital, outpatient clinic, and the emergency department (Appendix). The scenarios focused on material that students learn during their pre-clinical years such as chief complaint, present illness and past medical history. For the remainder of the session, about thirty minutes, students divided to groups of three, taking the roles of a Spanish-speaking patient, an English-speaking provider, and an interpreter to practice such real-life scenarios that one may encounter when providing healthcare. Although students took on the roles of “interpreters” during the scenarios, the goal of this was not to train them for such position. Rather, it was to provide the students with more practice of the phrases and improvement of comprehension and verbal Spanish skills that would be helpful when communicating with Spanish-speaking patients. The scenarios contained not only the relevant medical vocabulary, but also emphasized on culturally appropriate use of language when speaking to patients. Examples include using respectful phrases such as “ ¿Qué le trae por aquí?” which translates to “ what brings you here today?” rather than a literal translation of “ why are you here?” to “ ¿Por qué estás aquí?”, and always referring to patients as “ usted”, a formal form of “ you”, rather than “ tù”, the informal form. All sessions for both the beginner and intermediate group followed this agenda, however the vocabulary lessons and scenarios were more advanced for the intermediate group. At the end of the 5-week course, students took a post-course examination, with a format identical to the pre-course exam, to determine if there had been an improvement in Spanish language proficiency and the degree to which participants acquired Spanish language knowledge and skills. Statistical significance was determined using a *p*-value < 0.05. Students and faculty were not compensated or incentivized otherwise for their study participation.

## Results

Seven students, all members of the intermediate group, completed the course. Attendance among this group was 77.4%. In the self-assessment survey, all students reported that the style of role-play was beneficial to their learning and believed that the course helped improve their conversational Spanish skills. The majority of students, 85.6%, said this course helped improve knowledge of medical Spanish, while 71.4% of students believed the course helped them with becoming more culturally competent when providing care to Spanish-speaking patients.

Due to the non-normal distribution of data and small sample size, Wilcoxon signed-rank test was used for statistical analysis to compare repeated measurements before and after course completion on the same sample. When comparing pre-course to post-course examination scores, there was improvement of scores in all categories (Table 1). In particular, we found statistically significant improvement in oral translation of phrases from Spanish to English (*p*-value= 0.03)

**Table 1).**
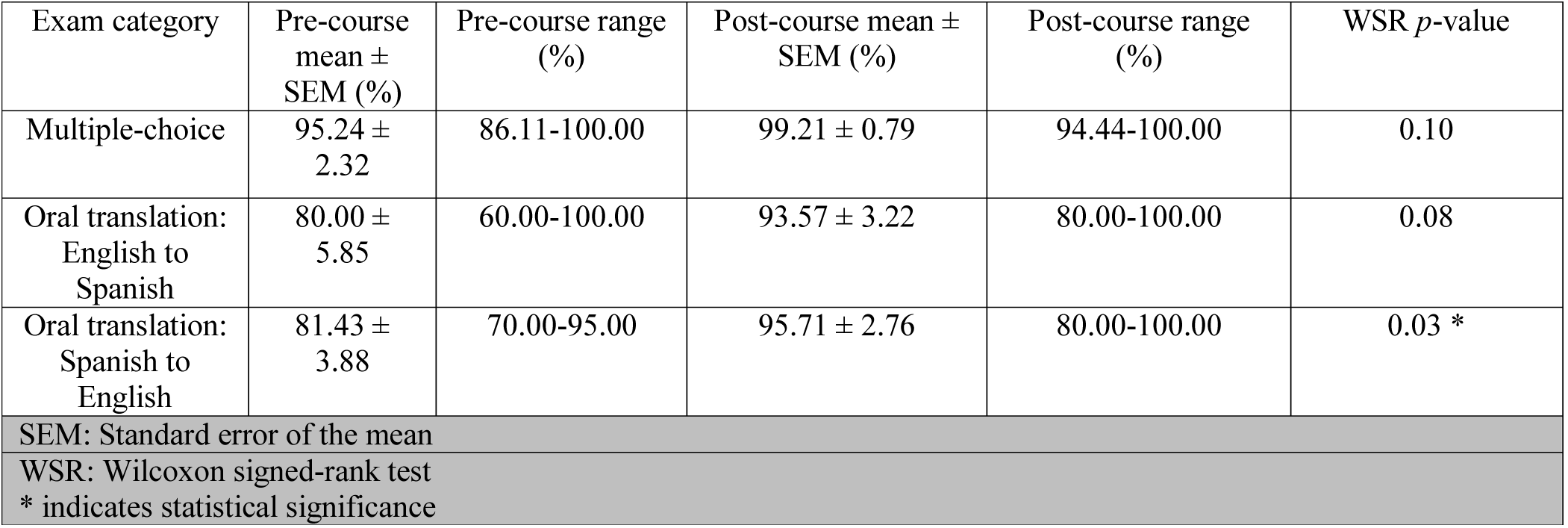
Comparison of pre-course and post-course examination scores in the intermediate group

## Discussion

Spanish is the most widely spoken non-English language in the United States [27]. As the number of Hispanic patients rises, the healthcare system faces the need for adequately skilled Spanish-speaking providers to deliver care to patients. For many years, health institutions have implemented medical Spanish classes in the attempt to bring forth such providers. However, many barriers to introducing effective courses have been identified [2,11,12]. In our study, we introduced a course that included components that Ortega et al., 2019 deemed important with respect to developing a comprehensive course [14]. These included involving other departments of the institution in designing such course and creating pre-course and post-course assessments to evaluate it. The pre-course assessment also served the purpose of dividing the students into two groups to ensure a more homogenous breadth of knowledge amongst them as too much heterogeneity in students’ language skills has shown to be a barrier in implementing medical Spanish courses [2]. We also incorporated important cultural nuances that one should take into account when providing care to Spanish-speaking patients, as these are equally as important as having the knowledge of the medical terminology and conversational skills [21–23]. The assessments also enabled us to objectively evaluate the knowledge of students as this has been a barrier to designing effective curricula [12,14–16]. In addition, the majority of the sessions’ time was focused on improving oral conversational skills, as oral proficiency is deemed most important when considering medical Spanish courses [24]. More importantly, achieving statistically significant improvement of Spanish comprehension was accomplished through a very minimally time requirement of one hour per week. This can be promising as lack of time was the most commonly reported barrier (51%) by medical schools in a national survey [2]. Additionally, this same national survey of medical school Spanish curricula reported that teaching modalities were largely varied with only 67% of programs utilizing student-to-student role play, as compared to 90% employing didactic instruction [2].

Our findings align with previous studies examining role-play. Nestel & Tierney, 2007 reported 96.5% (n=274) of students believed role-play was helpful for learning, which is closely mirrored by our finding that 100% of participants reported role-play was beneficial to their learning. We see this as an opportunity for increased up-take of role-play as a teaching modality.

Although the findings in our study provided promising results, this study had limitations of its own. The attrition rate among participants was 63% (12/19). Six of those students identified lack of time as the reason for withdrawing from the course. This highlights the recurring theme of lack of time as a notable obstacle to carrying out such courses. In addition, all five students in the beginner class withdrew from the course. Two students identified time as the reason while the other three stated to have felt that their baseline Spanish language skills were insufficient. Such contrast of background knowledge amongst the beginner and intermediate group is evident as the average number of years of completion of Spanish courses (high school and undergraduate education) for the intermediate participants was 6.5 years, while this number for the beginner students was 2.1 years. This suggests that perhaps some foundational knowledge of the language is needed in order for students to gain the most benefit from these courses. Given this course predominately focused on language production, further research is required to better understand how the role-play teaching modality benefits beginner students as compared to intermediate students. Special care should be taken when designing medical Spanish courses to address the interplay of baseline Spanish language skills and disciplinary approaches to teaching. In addition, the sample size that completed the course was only seven students, which is considerably small. This limited our statistical power and even though our results in all three categories of post-course assessment trended towards statistically significant findings, we were only able to yield such results in one category (Table 1). Although the assessments were objective and the content had been designed by faculty of the Spanish department, we did not utilize standardized patients for the assessments, as this has been the primary standard assessment of language skills across US medical schools [28]. We also evaluated the students’ knowledge only one week after completion of the course; therefore, our results do not provide insight on students’ ability to retain this knowledge long-term.

In summary, we investigated the potential benefit of a multidisciplinary approach with an emphasis on utilizing role-play as the central teaching method in a medical Spanish course. Intermediate level students demonstrated significant improvement with oral translation of phrases from Spanish to English. More importantly, this was accomplished through a minimal time requirement of one hour per week, as limited time poses a notable barrier to implementing a medical Spanish curriculum. Such findings call for further evaluation of the role-play teaching approach in a larger sample population, and perhaps formally evaluating the students in a setting of standardized patient interaction. We believe our study provides unique insight to the growing body of literature in developing effective courses to teach medical Spanish with a special emphasis on the under-utilized yet promising modality of role-play.

## Supporting information

Appendix: Spanish Course Materials

## Data Availability

N/A

## Acknowledgements

The authors thank Diego Zegarra for his early contribution to this project.

## References

1. Alonzo F. Hispanic Heritage Month 2019. 2019 [cited 2020 Jun 25]; Available from: https://www.census.gov/newsroom/facts-for-features/2019/hispanic-heritage-month.html

2. Morales R, Rodriguez L, Singh A, Stratta E, Mendoza L, Valerio MA, et al. National Survey of Medical Spanish Curriculum in U.S. Medical Schools. J Gen Intern Med. 2015;

3. Aitken G. Medical students as certified interpreters. AMA J Ethics. 2019;

4. Smedley BD, Stith AY, Nelson AR. Unequal Treatment: Confronting Racial and Ethnic Disparities in Health Care (with CD). Unequal Treat. Confronting Racial Ethn. Disparities Heal. Care (with CD). 2003.

5. Allison A, Hardin K. Missed Opportunities to Build Rapport: A Pragmalinguistic Analysis of Interpreted Medical Conversations with Spanish-Speaking Patients. Health Commun. 2020;

6. Ngo-Metzger Q, Sorkin DH, Phillips RS, Greenfield S, Massagli MP, Clarridge B, et al. Providing high-quality care for limited english proficient patients: The importance of language concordance and interpreter use. J Gen Intern Med. 2007;

7. Diamond L, Izquierdo K, Canfield D, Matsoukas K, Gany F. A Systematic Review of the Impact of Patient–Physician Non-English Language Concordance on Quality of Care and Outcomes. J. Gen. Intern. Med. 2019.

8. Ortega P, Pérez N, Robles B, Turmelle Y, Acosta D. Strategies for Teaching Linguistic Preparedness for Physicians: Medical Spanish and Global Linguistic Competence in Undergraduate Medical Education. Heal. Equity. 2019.

9. Wilson E, Chen AH, Grumbach K, Wang F, Fernandez A. Effects of limited English proficiency and physician language on health care comprehension. J. Gen. Intern. Med. 2005.

10. Schenker Y, Karter AJ, Schillinger D, Warton EM, Adler NE, Moffet HH, et al. The impact of limited English proficiency and physician language concordance on reports of clinical interactions among patients with diabetes: The DISTANCE study. Patient Educ Couns. 2010;

11. Mueller R. Development and evaluation of an intermediate-level elective course on medical Spanish for pharmacy students. Curr Pharm Teach Learn. 2017;

12. Reuland DS, Frasier PY, Slatt LM, Alemán MA. A longitudinal medical spanish program at one US medical school. J Gen Intern Med. 2008;

13. Fernández A, Pérez-Stable EJ. ¿Doctor, habla español? Increasing the Supply and Quality of Language-Concordant Physicians for Spanish-Speaking Patients. J Gen Intern Med. 2015;

14. Ortega P, Pérez N, Robles B, Turmelle Y, Acosta D. Teaching Medical Spanish to Improve Population Health: Evidence for Incorporating Language Education and Assessment in U.S. Medical Schools. Heal. Equity. 2019.

15. Ortega P. Spanish language concordance in U.S. Medical care: A multifaceted challenge and call to action. Acad Med. 2018;

16. Diamond LC, Reuland DS. Describing physician language fluency deconstructing medical spanish. JAMA - J. Am. Med. Assoc. 2009.

17. Diamond LC, Schenker Y, Curry L, Bradley EH, Fernandez A. Getting By: Underuse of interpreters by resident physicians. J Gen Intern Med. 2009;

18. Ortega P, Diamond L, Alemán MA, Fatás-Cabeza J, Magaña D, Pazo V, et al. Medical Spanish Standardization in U.S. Medical Schools: Consensus Statement From a Multidisciplinary Expert Panel. Acad Med. 2020;

19. Burbano O’Leary SC, Federico S, Hampers LC. The truth about language barriers: one residency program’s experience. Pediatrics. 2003;

20. Hardin K. An overview of medical Spanish curricula in the United States. Hispania. 2015;

21. Lopez L, Vranceanu AM, Cohen AP, Betancourt J, Weissman JS. Personal characteristics associated with resident physicians’ self perceptions of preparedness to deliver cross-cultural care. J Gen Intern Med. 2008;

22. Haskard Zolnierek KB, Dimatteo MR. Physician communication and patient adherence to treatment: A meta-analysis. Med Care. 2009;

23. Ghaddar S, Ronnau J, Saladin SP, Martínez G. Innovative approaches to promote a culturally competent, diverse health care workforce in an institution serving hispanic students. Acad Med. 2013;

24. Hardin KJ, Hardin DM. Medical Spanish Programs in the United States: A Critical Review of Published Studies and a Proposal of Best Practices. Teach Learn Med. 2013;

25. Nestel D, Tierney T. Role-play for medical students learning about communication: Guidelines for maximising benefits. BMC Med Educ. 2007;

26. Jackson VA, Back AL. Teaching communication skills using role-play: An experience-based guide for educators. J Palliat Med. 2011;

27. The 2020 Census Speaks More Languages [Internet]. [cited 2020 Jun 25]. Available from: https://www.census.gov/newsroom/press-releases/2020/languages.html

28. Karkowsky CE, Chazotte C. Simulation: Improving communication with patients. Semin. Perinatol. 2013.

