## Appendix: Spanish Course Materials for "A Multidisciplinary Approach: Teaching Medical Spanish to Medical Students Using Role-play"

### Pre-course examination
### multiple choice

**1. mi hermano tuvo una piedra cuando tenía treinta años.**

a) My father had an infection when he was forty years old
b) My brother had a stone when he was thirty *
c) My cousin had cancer when he was fifty
**d)** My uncle had an infection when he was sixty

**2. the results have arrived, and the CT scan showed us that you have a stone**

a) Los resultados han llegado y la tomografía computarizada enseñó que usted tiene una piedra *
b) Los resultados han llegado y la tomografía computarizada enseñó que usted tiene una masa
c) Los datos dicen que la piedra está en la tomografía computarizada
d) Los rayos-X enseñan que hay una piedra con infección

**3. Me duele mucho la espalda, este dolor es insoportable. Hace dos horas que empezó y ya no lo puedo aguantar.**

a) My back hurts very much, this pain is unbearable. It started yesterday and I can't tolerate it
b) My shoulder hurts very much, this pain is bad. It started today and I can't tolerate it
c) My back hurts very much, this pain is unbearable. It started two hours ago, and I can't tolerate it *
d) I have a bad headache and I can't take it anymore.

**4. Mi numero de teléfono es doscientos treinta y ocho- setenta y cinco, veinticuatro**

a) My phone number is 238-75-24 *
b) My phone number is 939-71-22
c) My phone number is 231-33-21
d) My phone number is 674-31-98

**5. Hace un año que perdí mi seguro médico.**

a) I lost my job last year
b) I lost my health insurance last year *
c) She lost her job last year
d) I lost my son last year

**6. Can you please tell me if you have had a fever and other symptoms?**

a) ¿Dígame por favor si ha tenido fiebre y otros síntomas? *
b) ¿Dígame si tiene fiebre y que le trae por aquí?
c) ¿Ha tenido fiebre y vómitos?
d) ¿Dígame por favor su fecha de nacimiento?

"*" Indicates the correct answer

### **Pre-course examination**
### **oral translation**

Translate the following phrases from Spanish to English or from English to Spanish

Nurse: Can you please confirm your date of birth and your phone number?
Patient: Hola, 22 de octubre de 1939. Mi número es: 205-981-1366.

Nurse: Thank you for the information. I am going to speak to the doctor and come back shortly
Patient: Me parece bien, gracias a usted.

Nurse: Have you taken any medications to help with the pain?
Patient: Sí, tomé dos pastillas, pero no se me ha quitado este dolor.

Nurse: Can you please tell me if you have had a fever and have had other symptoms?
Patient: No, no he tenido fiebre, pero me siento débil, mareado, y tengo náusea.

Nurse: Who do you live with?
Patient: Con mis hijos, mi esposa y mi suegra.

### Intermediate group practice scenario
### En el departamento de emergencia

Enfermera: Good afternoon, what brings you here?

Paciente: Buenas tardes. Me duele mucho la espalda, este dolor es insoportable. Hace dos horas que empezó y ya no lo puedo aguantar.

Enfermera: Tell me more please!

Paciente: Fui a mi trabajo esta mañana y estaba bien. Llegué a mi casa a las cuatro de la tarde y tomé una siesta. Desperté a las 4:30 porque sentía algo raro en el estómago y en la espalda. De repente, sentí un dolor muy fuerte en mi lado izquierdo. El dolor no se me quitaba y también vomité 2 veces. Cuando mis hijos llegaron, les dije lo que pasó y me trajeron al hospital.

Enfermera: Have you taken any medications to help with the pain?

Paciente: Sí, tomé dos pastillas, pero no se me ha quitado este dolor. ¡Por favor, ayúdeme señora, ya no lo aguanto!

Enfermera: Soon we are going to do some studies to find out the cause of your pain. Tell me more, have you had kidney stones?
Paciente: No, solo he tenido una cirugía en toda mi vida. Me sacaron la vesícula hace casi 5 años.

Enfermera: Anyone in your family has had kidney stones: parents, brothers, cousins?
Paciente: Sí, mi hermano tuvo una piedra cuando tenía treinta años. Mi tía también, pero no sé cuántos años tenía.

Enfermera: Thank you for the information. We are now going to take a CT scan to see if you have a stone in your left kidney. We are also going to get a urine sample.
Paciente: Está bien, gracias.

Una hora después…

Enfermera: Hello, the results have arrived, and the CT scan showed us that you have a stone and there is blood in your urine, but we believe that the stone will pass, and you can go home with medication to calm the pain.

Paciente: Gracias, ahora me siento mejor.

### Beginner group practice scenario

#### En el hospital

Nurse: Good morning Mr. Lopez, How are you today?
Mr. López: Buenos días. Estoy bien, gracias.

Nurse: Do you want to have food?
Mr. López: Sí, tengo mucha hambre.

Nurse: Do you want to drink water?
Mr. López: Sí, tengo mucha sed.

Nurse: Is your wife here?
Mr. López: Sí, ella está aquí.

Nurse: Do you want to walk today?
Mr. López: Ahora no, estoy muy cansado.

Nurse: That is alright, but you do have to walk in the afternoon.
Mr. López: Claro, voy a caminar con mi esposa por la tarde.

Nurse: Do you have pain right now?
Mr. López: Sí, me duele la espalda.

Nurse: I am going to come back in 10 minutes with your medications.
Mr. López: Perfecto, hasta luego.
